# Genetic variants underpinning lung function decline in the Lifelines general population cohort study

**DOI:** 10.1101/2025.10.26.25338838

**Authors:** Rui Marçalo, Corry-Anke Brandsma, Peter J. van der Most, Alda Marques, Gabriela R. Moura, Maarten van den Berge, Judith M Vonk, Maaike de Vries

## Abstract

**Rationale:** Variations in age-related lung function decline are associated with genetic and environmental factors. The genetic variants contributing to this decline remain largely unknown, limiting the understanding of individual susceptibility and potential interventions.

**Objectives:** This study aims to uncover the genetic factors associated with lung function decline, using the Lifelines cohort study.

**Methods:** Longitudinal data covering 3 visits and approximately 15 years follow up from 165,138 individuals were available. Genotyping and longitudinal spirometry data were present for 24,749 subjects aged 25 or older at baseline. A three-step approach was used. First, lung function change over time was estimated using a linear-mixed effect model. Second, a genome-wide association study for the estimated change was conducted, adjusting for age, sex, smoking load and the first 10 principal components. Third, single nucleotide polymorphisms (SNPs) with p-value<1×10^−5^ were analysed using a linear-mixed effect model. Selected SNPs were tested in two independent cohorts (N=1,376 and N=27,249), followed by meta-analysis.

**Measurements and Main Results:** Among the included individuals, 19,722 had spirometry at two timepoints and 5,027 at three, with median [IQR] follow-up of 8.1 [4.4, 10.4] and 12.1 [11, 13.6] years, respectively. We identified 67 variants suggestively associated with lung function decline (p<1×10^?5^). Rs150094594, was significant (P=0.007) in a replication cohort with consistent effect direction. This intergenic variant, not previously reported, reached genome-wide significance in the meta-analysis (rs150094594:T, β(SE): 15.9(2.7) ml/year, p-value=3.9×10^−9^).

**Conclusions:** This study shows a role of genetics in lung function decline, emphasizing the importance of exploring the interplay between genetic and environmental factors.

## Introduction

Lung function follows a predictable trajectory, with growth and development reaching its peak in early adulthood, followed by a steady decline during the remaining lifespan [1, 2]. The most common method for assessing lung function is through spirometry, where forced expiratory volume in the first second (FEV1) and forced vital capacity (FVC) are measured, providing information about the health of the airways and lungs. For most individuals, FEV1 decline remains within a range that does not result in clinical symptoms [1]. However, for some individuals, lung function declines more rapidly than normal. This accelerated lung function decline increases the risk of developing chronic obstructive pulmonary disease (COPD), a leading cause of mortality and morbidity worldwide [1, 3].

The aetiology of accelerated lung function decline remains poorly understood. Although environmental factors (e.g., smoking, air pollution and smoke from indoor cooking) are thought to be the key contributors, they do not fully explain the variability observed among individuals [1, 4]. Approximately 30% of smokers maintain normal lung function throughout their lives [5], thus, suggesting that other factors, including genetic predisposition, may play an important role in lung function decline [6]. Genome-wide association studies (GWAS) have identified over 1000 genetic variants associated with lung function [7, 8]. However, the majority of GWASs have been performed on cross-sectional studies, with the FEV1 measurement reflecting the combination of an individual’s maximal attained lung function in early adulthood and the decline up to that specific moment. As such, these studies have conflated genetic factors affecting maximal attained lung function with those influencing the rate of decline [8, 9], making it challenging to disentangle the aetiologies of these two mechanisms that contribute to COPD. Moreover, the complexity and multifactorial aspect of lung function decline coupled with the limited availability of large longitudinal studies on lung function have hindered the robustness and replicability of the findings [7].

With this study, we aim to identify genetic variants associated with lung function decline in the general population using the Lifelines cohort study, a large population-based multi-generation cohort.

## Methods

### Study population

This study was conducted using data from the Lifelines cohort study [10, 11]. Briefly, Lifelines is a multi-disciplinary prospective population-based cohort study examining in a unique three-generation design the health and health-related behaviours of 167,729 persons living in the North of the Netherlands. It employs a broad range of investigative procedures in assessing the biomedical, socio-demographic, behavioural, physical and psychological factors which contribute to the health and disease of the general population, with a special focus on multi-morbidity and complex genetics. Sociodemographic (i.e., age, sex), anthropometric (i.e., weight, height), and clinical data (i.e., smoking status and load, lung function), as well as biological samples were collected through structured questionnaires, physical measurements and laboratorial analyses [10, 11]. Currently, data is available from three visits with approximately 5 years between the visits. The Medical Ethics committee from the University Medical Center Groningen approved Lifelines (UMCG METc 2007/152), and written informed consent was collected from all participants.

### Lung function (decline)

For this study, individuals aged 25 years or older at baseline, with at least two lung function measurements (to compute change in lung function over time), and with available genotype data were selected. Pre-bronchodilator spirometry was performed with a Welch Allyn Version 1.6.0.489, PC-based Spiroperfect with CardioPerfect workstation software according to established guidelines [12]. Results and technical quality were evaluated by well-trained assistants and difficult to interpret results were re-evaluated by a lung physician. FEV1 in milliliters was used to study lung function decline.

### Genotyping

Genotyping was performed in three batches using the Illumina CytoSNP-12 Bead Chip v2 array (CytoSNP), Infinium Global Screening Array Beadchip-24 v1.0 (GSA) and FinnGen Thermo Fisher Axiom custom array (Affy) for batch 1, 2 and 3, respectively. Low quality samples (genotyping rate below 80%) and markers (call rate below 80%) were excluded through a two-step procedure of call rate thresholding. Genotyping errors were assessed by examining samples’ heterozygosity (>4 standard deviations) and markers’ deviation from Hardy-Weinberg equilibrium (*P*<1×10^−06^). Concordance between self-reported and genotyped sex was evaluated. Moreover, population stratification was inspected by principal component analysis and rare variants (minor allele frequency < 1%) were excluded [13]. All three arrays were imputed against the Haplotype Reference Consortium [14]. After quality control, there were 17,517,757 (CytoSNP), 18,810,061 (Affy), and 18,682,597 (GSA) single nucleotide polymorphisms (SNPs) available.

### Statistical analyses

A three-step approach was employed to perform a GWAS on lung function change over time in Lifelines within the available computational power.

First, lung function change over time was estimated for each subject using a linear mixed effect model (LME) with pre-bronchodilator FEV1 (mL) as primary outcome and time (defined as the number of years between the FEV1 measurement and the baseline assessment) as independent variable. The LME incorporated a random intercept and a random effect for time to account for individual variability in both baseline FEV1 and in change over time. Additionally, a fixed effect for time was included. For each subject, the estimated lung function change was calculated as the sum of the fixed and the random effect for time. A positive estimated lung function change indicated a gain in FEV1 over time, while a negative estimated lung function change indicated a decline of FEV1 over time. Estimation was performed using the *nlme* package from R software (version 4.2.1.) [15].

Second, GWASs were conducted per genotyping array (CytoSNP, Affy and GSA) using the estimated lung function change (mL/year) as outcome, and adjusted for age, sex, smoking load (pack-years), and the first 10 principal components to adjust for genetic ancestry. GWASs were performed using REGENIE (version 3.2.2), to account for sample relatedness within each genotyping array [16–18]. If a sample had close relatives genotyped on a different genotyping array, only one member of the group of relatives remained included in the analysis, to ensure the necessary data independence for the combination of GWASs. Priority was given to participants genotyped with GSA over Affy/CytoSNP, and Affy over CytoSNP. The results from the three GWASs (CytoSNP, Affy, GSA) were combined through a meta-analysis using the software METAL [19]. SNPs with *P* < 1×10^−5^ were selected for external replication.

Finally, to obtain the most accurate estimates, we performed an LME on the SNPs identified in the previous step, as this was now computationally feasible. The model included time, SNP and their interaction as fixed effects, along with random effects for both the intercept and slope (i.e., time). Modelling was performed using the *nlme* package from R software (version 4.2.1.) [15].

### Replication

Replication was performed using data from 1) the Vlagtwedde-Vlaardingen (Vla-Vla) cohort and 2) CHARGE-SpiroMeta cohort. [7, 20].

The Vla-Vla cohort is a general population-based cohort of Dutch individuals who were followed longitudinally every 3 years over a 25-year period. At each follow-up visit, sociodemographic, anthropometric, and respiratory symptoms data were collected [21]. Additionally, a blood sample was taken and spirometry was performed [21, 22]. Comprehensive details on DNA extraction, genotyping method, quality control protocol and imputation procedure for this cohort have been reported previously [21]. Lung function change over time was modelled using an LME with random intercepts and slopes. Fixed effects included time (defined similarly to Lifelines), the SNP of interest, and their interaction. The modelling was performed using the *nlme* package from R software (version 4.2.1.) [15, 23]. When a selected SNP was not available in the Vla-Vla cohort, a SNP with a linkage disequilibrium (LD) r^2^ > 0.8 was used as proxy. If multiple proxy SNPs were available, the SNP with the highest r^2^ was chosen.

The replication on the CHARGE-SpiroMeta cohort was performed by leveraging the summary statistics of their published GWAS on longitudinal change in adult lung function, that included 14 cohorts of European ancestry [7]. Detailed description of the study design and cohorts included are available elsewhere [7]. The SNPs identified in Lifelines were extracted from the summary statistics of the CHARGE-SpiroMeta cohort’s GWAS, followed by an evaluation of its significance and effect. SNPs with a P < 0.05 and same direction of effect as in Lifelines were considered replicated. When a selected SNP was not present, a proxy SNP with r^2^ > 0.8 was extracted. If multiple proxy SNPs were available, the SNP with the highest r^2^ was chosen.

### Meta-analysis

A meta-analysis including Lifelines, Vla-Vla and CHARGE-SpiroMeta cohorts was conducted to determine the combined effect size of the SNPs. The meta-analysis was performed only for the subset of SNPs initially identified as associated with lung function change in Lifelines, using METAL [19].

## Results

### Lifelines cohort characterisation

Of the initial 165,138 participants, 50,305 were eligible adults over 25 years old with two or three FEV1 measurements available. Of these, 22,952 had no genotype data available and 2,604 were related to other study participants genotyped using a different genotyping array, and were therefore excluded. A total of 24,749 participants were included in this study, distributed across genotyping arrays as follows: CytoSNP (7,280), Affy (7,723), and GSA (9,746) (Figure 1).

**Figure 1.**
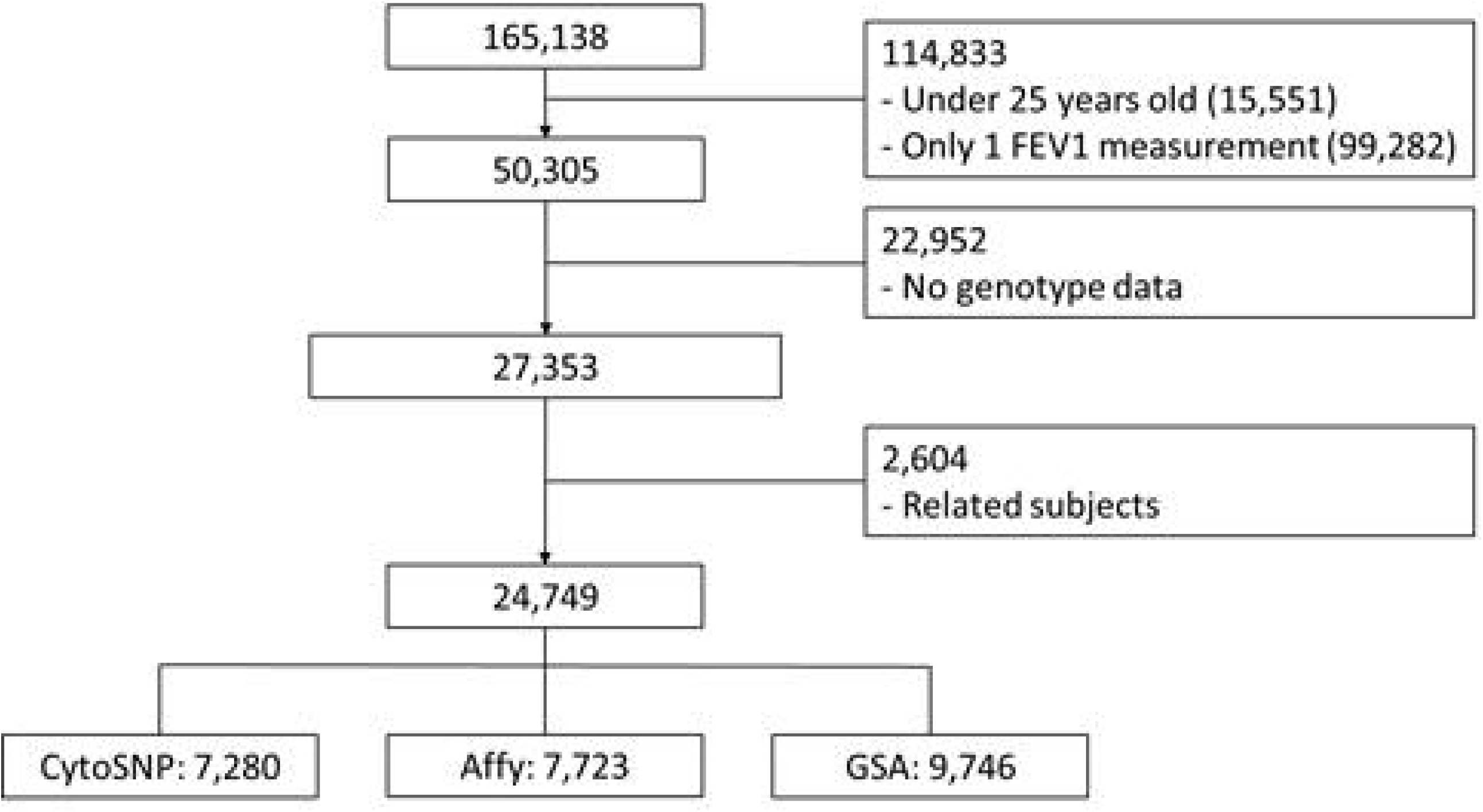
Selection flow chart of Lifelines cohort study participants.

At baseline, the study population was predominantly female (59.9%), with a median [IQR] age of 44 [37, 51] years and BMI of 25.4 [23.2, 28.1] Kg/m^2^. Most participants were either never smokers (45.4%) or former smokers (35.6%) (Table 1). Of the included individuals, 5,027 had three timepoints with lung function measurements available with a follow-up period of 12.1 [10.9, 13.6] years. The remaining 19,722 had only two timepoints with lung function measurements, with a follow-up period of 8.1 [4.4, 10.4] years.

**Table 1.** Baseline characteristics of Lifelines cohort study (N=24,749).

|  | Lifelines cohort (N=24,749) |
| --- | --- |
| Age, years (median [IQR]) | 44.0 [37, 51] |
| Sex, female n (%) | 14,822 (59.9%) |
| BMI, Kg/m<sup>2</sup> (median [IQR]) | 25.4 [23.2, 28.1] |
| Smoking status, n (%) |  |
| Never smoker | 11,115 (45.4%) |
| Former smoker | 8,722 (35.6%) |
| Current smoker | 4,655 (19.0%) |
| Smoking load, pack-years (median [IQR]) | 8.3 [3.5, 15.5] |
| FEV<sub>1</sub>, liter (median [IQR]) | 3.4 [2.9, 4.1] |
| FEV<sub>1</sub>, % of predicted (median [IQR]) | 96.8 [88.7, 105] |
| FEV<sub>1</sub>/FVC (median [IQR]) | 77.4 [73.1, 81.4] |
| FEV<sub>1</sub> change over time, mL/year (mean ± SD) | -24.2 ± 10.8 |

*Description: BMI: body mass index; FEV<sub>1</sub>: forced expiratory volume in one second; FVC: forced vital capacity.*

### Discovery meta-analysis

The GWASs conducted on the Lifelines cohort did not reveal any genome-wide significant SNPs (*P*<5×10^−8^), while 67 SNPs across 32 independent loci were suggestively associated with lung function decline (*P*<1×10^−5^) (Supplementary table 1). To obtain more precise effect estimates, an LME model was applied to the 67 SNPs (Table 2). Among the associated SNPs, the largest effects were observed for rs150094594 (chr7:62022453:T, β (SE): 15.4 (2.9) mL/year), rs62066418 (chr17:4481591:A, β (SE): 11.5 (2.5) mL/year) and rs117883345 (chr7:62210855:T, β (SE): 10.8 (2.9) mL/year), all with effect sizes above 10 mL/year. For all three SNPs, the major allele was associated with a faster lung function decline, and their allelic frequencies were consistent with the reported frequencies for European populations.

### Validation

Individuals from the Vla-Vla cohort (N=1,376) presented an age of 33 [27, 40] years old at baseline and were mainly female (53%). Participants were followed for a median of 7 [5, 8] visits over 19 [15, 22] years. The SNPs rs150094594 and rs12501883 demonstrated nominal significance, of which only rs150094594 showed the same direction of effect as in Lifelines (rs150094594:T, β (SE): 19.4 (7.2) mL/year, *P*=0.007; and rs12501883:A, β (SE): −1.5 (0.7) mL/year, *P*=0.04) (Supplementary table 2).

In the CHARGE-SpiroMeta cohort, rs4820911 and rs11030451 were nominally significant but both with an effect in the opposite direction of Lifelines (rs4820911:G, β (SE): −14.3 (7.2) mL/year, *P*=0.047; and rs11030451:C, β (SE): 7.4 (3.6) mL/year, *P*=0.04) (Supplementary table 2).

### Meta-analysis

The 67 SNPs identified in Lifelines as suggestively associated with lung function decline were meta-analysed with the Vla-Vla and CHARGE-SpiroMeta cohorts. Among them, 46 were present in both validation cohorts, 20 were present in only one of them, and one was not present in neither replication cohort. Four SNPs (rs62066418, rs36022992, rs113849236, rs12187807) reached the suggestiveness threshold (*P* < 1 x 10^−5^) and one reached genome-wide significance (rs150094594:T, β (SE): 15.9 (2.7) mL/year, *P*=3.9 x 10^−9^) (Figure 2 and supplementary table 2). In all five SNPs, the minor allele showed a positive β-coefficient, being therefore associated with a slower lung function decline (Figure 2 and supplementary table 2).

**Figure 2.**
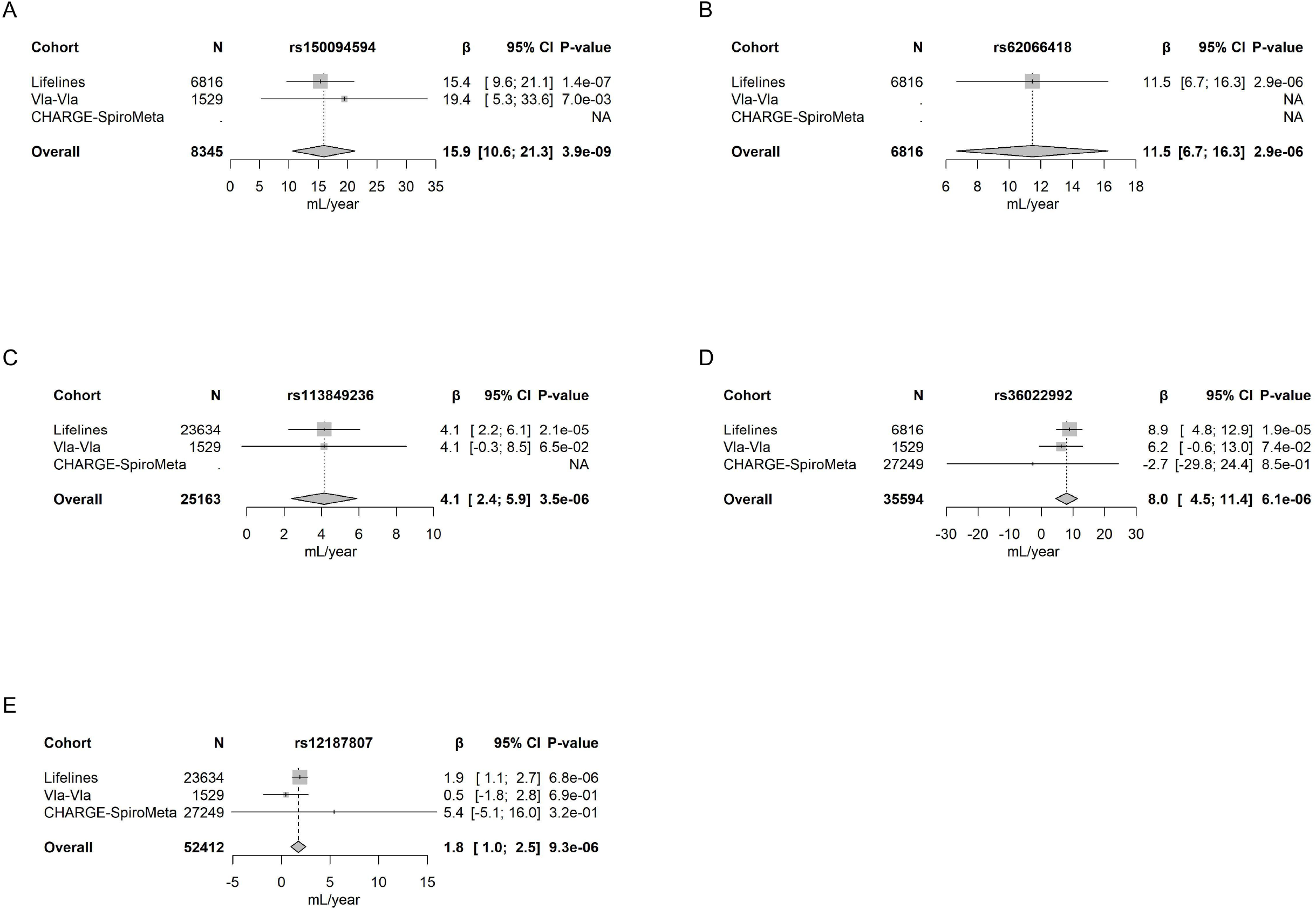
Forest plot of SNPs associated with lung function decline in the meta-analysis of Lifelines cohort study, Vlagtwedde-Vlaardingen cohort and the CHARGE-SpiroMeta cohort. A) rs150094594; B) rs62066418; C) rs113849236; D) rs36022992; E) rs12187807. N: sample size; β: beta-coefficient; 95% CI: confidence interval.

## Discussion

Within the Lifelines discovery cohort, consisting of over 24,000 individuals, we identified 67 SNPs suggestively associated with lung function change over time. Among them, rs150094594 showed an effect estimate of 15.4 mL/year in the discovery cohort and was successfully replicated in one of the two independent replication cohorts, where it exhibited an effect estimate of 19.4 mL/year. The 67 SNPs identified in the discovery cohort were meta-analysed alongside the two independent cohorts. The SNP rs150094594 reached genome-wide significance while four other SNPs retained a suggestive p-value.

Lifelines is a population-based cohort, and both FEV1 at baseline (3.4 L) and change over time (−24.2 mL/year) were consistent with what has been reported previously (FEV1: 3.9 L and FEV1 change: −29.2 mL/year) [7, 24]. In the discovery GWAS, a “3-step approach” was used, first estimating the FEV1 change in mL/year for each subject, then performing a GWAS on the estimated change, and finally running a full LME including both change over time and the effect of the SNP on this change for the selected SNPs. This approach was taken to overcome computational power constraints inherent to performing an LME for approximately 18 million SNPs in the dataset. Under the reported assumption that lung function, after reaching its peak in early adulthood, follows a steady and linear decline, our approach seems valid and is supported by the literature [1, 2, 4]. However, with first estimating the FEV1 change in ml/year per individual and then analyzing the association between each SNP and this estimated change, we do not simultaneously model both within- and between-individual differences. This approach is therefore less powerful than performing a full LME including both change over time and the effect of the SNP on this change in one model. For this reason, after combining the results from the GWASs on the estimated change over time on CytoSNP, Affy and GSA, the effect estimates for the identified SNPs were recalculated using a full LME. The SNPs’ significance levels were consistent between the two approaches (data not shown), proving this to be a valid option for future research studies where the computational resources required to run an LME on millions of SNPs are limited.

Four of the SNPs identified in Lifelines were nominally significant in at least one of the replication cohorts. However, of those four, only rs150094594 showed the same direction of effect in both discovery and replication cohorts, with meaningful estimates above 15 mL/year. The effect allele of rs150094594 (T) showed a very low and similar frequency between the Lifelines and Vla-Vla cohorts, around 1%, with a comparable effect on lung function change. Rs150094594 is an intergenic single nucleotide variation from C to T, located in chromosome 7 at position 62022453 (GRCh37), and does not have any other SNP in high LD that could be used as proxy when not present in a replication cohort (e.g., not available in the CHARGE-SpiroMeta cohort). An eQTL analysis was performed but yielded no association between rs150094594 and any gene (data not shown).Additionally, it has not been reported as associated with any disorder, with the closest gene being the pseudogene RNU6-417P. As such, it poses a difficult challenge in its biological interpretation and relevance.

The meta-analysis of the 67 SNPs identified in Lifelines, combining the three cohorts (Lifelines, Vla-Vla, and CHARGE-SpiroMeta cohorts), unravelled one significantly associated SNP and four SNPs suggestively associated with lung function change over time. As previously mentioned, rs150094594 has not been reported as associated with any condition, therefore precluding its biological framing. Two of the other four suggestively associated SNPs, rs62066418 and rs12187807, are also intergenic variants that have not been previously reported as linked to any condition or gene, impeding their biological interpretation. The SNP rs113849236 is an intronic variant of *SNAP47*, a gene that has been recently associated with picornaviruses infections. One genera of the picornavirus’s family are rhinoviruses, a type of virus associated with respiratory infections (e.g., common cold) [25–27]. *SNAP47* appears to be vital for the virus infection process, as *SNAP47* knock down has been shown to significantly reduce viral titer, though its mechanism of action has not been fully described [25, 26]. Exacerbations of chronic lung diseases such as COPD and asthma can commonly be triggered by viral infections [28, 29] and are major contributors to steep declines in lung function, usually not fully recoverable to the pre-exacerbation FEV1 level [30, 31]. Based on these premises, it seems reasonable to speculate that the rs113849236 C-allele, which was associated with a slower lung function decline (positive β-coefficient), might trigger a down-regulation of *SNAP47*, culminating in a less efficient infection process by viruses. Consequentially, it could make individuals less prone to exacerbations, protecting them from a steep drop in lung function. Additionally, *SNAP47* has also been associated with non-small cell lung cancer, but without a clear relation to lung function, besides sharing smoking as the main risk factor. [32, 33]. Finally, rs36022992 is an intronic variant of *MYO3B*, which has been linked with mucociliary clearance in cystic fibrosis [34]. Deficient mucociliary clearance, a pronounced feature of accelerated lung function decline-derived conditions (e.g., COPD), can lead to mucus retention and bacterial infection, increasing the risk of exacerbations and consequent FEV1 decline [34–36]. The effect of rs36022992 on *MYO3B* and its subsequent impact on mucociliary clearance is yet to be described. As such, no hypotheses or conclusions could be drawn at this stage. Finally, it is worth noting that four of the five SNPs identified in the cohort’s meta-analysis have a low minor allele frequency, below 2%. Low frequency (1-5%) and rare (<1%) variants are thought to play a substantial part in complex diseases [37, 38]. Despite this, they have been largely underexplored due to the challenges associated with their identification and replication. However, when identified, these variants often exhibit larger effect sizes compared to common variants, suggesting they may have a significant impact on disease susceptibility and progression [37, 38]. Overall, our results seem to suggest a moderate impact of genetics on lung function decline. From the initial GWAS on over 24,000 individuals, 67 SNPs emerged but none reached genome-wide significance. Moreover, five emerged from the meta-analysis, of which only one reached genome-wide significance. A recent GWAS (not yet peer reviewed) on 52,056 individuals from multiple ancestries has reported a similar number of SNPs associated with lung function decline [39], thus reinforcing that genetics alone carry a moderate weight in lung function decline [40].

## Limitations and future work

Our study has some limitations that should be acknowledged. Lifeline’s genotyping was performed using three different arrays (CytoSNP, Affy and GSA), with data being combined through a post-GWAS meta-analysis. While efforts are underway to synchronize all data, variations in genotyping arrays may introduce variability and certain SNPs may be missing or imputed with lower accuracy. Due to computational limitations, Lifeline’s initial SNP screening (over 24,000 people and 18,000,000 SNPs) used a three-step approach instead of a direct LME, with LMEs later performed on the selected SNPs for more accurate estimates. We assessed the differences in significance between approaches for the top 500 SNPs and found the p-values to be comparable, thus supporting our methodological option. Despite leveraging high LD SNPs as proxies, some of the SNPs could not be replicated due to not being present in the independent cohorts, highlighting the need for broader SNP coverage in future studies.

As future perspectives, functional validation of the SNPs identified in this study could provide deeper insights into their biological roles and relevance in lung function decline, paving the way for translational applications in personalised medicine. Additionally, research should prioritize gene-environment interaction (GxE) analyses to uncover how genetic variants modulate trait expression in response to environmental factors (e.g., smoking and air pollution). Certain SNPs may only exert their effects in the presence of these environmental triggers, and uncovering these interactions would further our understanding of these complex interplays.

## Conclusion

This study represents one of the largest GWAS investigating the genetic factors underpinning lung function decline. Among 67 SNPs suggestively identified in the discovery cohort, one replicated in an independent cohort, exhibiting an effect size greater than 15 ml/year. The meta-analysis combining three cohorts further confirmed the association of rs150094594 with lung function decline. Two of the other associated SNPs suggest genetic effects related to viral infection efficiency and mucociliary clearance, as possible contributors to lung function decline. Despite these findings, the non-replication of the majority of the SNPs in the combined meta-analysis suggests a moderate influence of genetic factors on this trait. This underscores the need of better characterized cohorts (e.g., occupational exposures) and highlights the growing importance of gene-environment interactions to better understand the complexities of lung function decline.

## Data Availability

All data produced in the present study are available upon reasonable request to the authors.

## Acknowledgments

The authors wish to acknowledge the services of the Lifelines Cohort Study, the contributing research centres delivering data to Lifelines, and all the study participants.

## Data availability statement

Data may be obtained from a third party and are not publicly available. Researchers can apply to use the Lifelines data used in this study. More information about how to request Lifelines data and the conditions of use can be found on their website.

